# Interpretable Deep Learning Approaches for Reliable GI Image Classification: A Study with the HyperKvasir Dataset

**DOI:** 10.1101/2025.07.22.25332009

**Authors:** Saif Bin Wahid, Zarin Tasnim Rothy, Raisul Kabir News, Sakib Anwar Rieyan

**Author notes:** Corresponding Author: Sakib Anwar Rieyan.

## Abstract

Deep learning has emerged as a promising tool for automating gastrointestinal (GI) disease diagnosis. However, multi-class GI disease classification remains underexplored. This study addresses this gap by presenting a framework that uses advanced models like InceptionNetV3 and ResNet50, combined with boosting algorithms (XGB, LGBM), to classify lower GI abnormalities. InceptionNetV3 with XGB achieved the best recall of 0.81 and an F1 score of 0.90. To assist clinicians in understanding model decisions, the Grad-CAM technique, a form of explainable AI, was employed to highlight the critical regions influencing predictions, fostering trust in these systems. This approach significantly improves both the accuracy and reliability of GI disease diagnosis.

## I. Introduction

Medical image analysis is crucial for diagnosing and treating gastrointestinal (GI) diseases, with AI playing an increasing role. Accurate classification of intestinal sections is necessary to detect abnormalities. Endoscopy and colonoscopy are common but complex image-based diagnostic methods. However, manual image interpretation is time-consuming, prone to errors, and can delay diagnosis, especially when further tests like biopsies are needed. Deep learning has improved automated image classification, helping doctors make quicker, more accurate decisions. Still, classifying intestinal images remains difficult due to the complexity and variability of the anatomy and disease features [15] [16].

The main problem in classifying gastrointestinal images is the lack of reliable automated systems that can accurately identify different parts of the intestine. The variation in textures, shapes, and lighting across images makes it difficult for current models to work well with new data. While many deep learning models have been tried, they still lack the precision needed for clinical use and are often not easy to understand, which reduces doctors’ trust in them [15] [16].

This research seeks to enhance the accuracy and transparency of models through the application of explainable AI (XAI) methods, such as Grad-CAM. By increasing the models’ transparency, we aim to foster trust and improve their applicability in diagnosing conditions like Crohn’s disease, ulcerative colitis, and colorectal cancer [15] [16]. The central research question explored in this study is: *How can we improve the accuracy and interpretability of automated classification models for various sections of the intestine by employing deep learning and explainable AI techniques?*

This research adds to the field of medical image analysis in the following ways:

1. This research classifies different sections of the intestine using the HyperKvasir dataset by applying advanced deep learning models like VGG19, ResNet, Dense Net, Efficient Net, and Inception Net.

2. Several boosting techniques, which merge multiple weak learners to create a stronger predictive model, were employed to enhance the accuracy and robustness of the AI model for clinical applications.

3. Grad-CAM (Gradient-weighted Class Activation Mapping) was utilized to increase the model’s interpretability by emphasizing key areas in the images that impacted its decisions, thereby promoting transparency and trust in clinical settings.

## II. Literature Review

Gastrointestinal diseases can lead to cancer, making early detection crucial. Advances in deep learning and machine learning are enabling computer-assisted diagnosis systems to efficiently classify gastrointestinal images. This technology supports gastroenterologists in diagnosing and treating conditions more effectively, addressing the challenges of manual diagnosis.

To facilitate the process Borgli et al. [1] conducted research by making use of the HyperKvasir dataset, which contains 10662 labeled images of the both upper and lower gastrointestinal tract, to demonstrate the potential benefits by using ML techniques for identifying, and detecting and examine gastrointestinal abnormalities and obtained a precision of 0.633 and an MCC of 0.902 by combining ResNet-152 and DenseNet-161. Later on, Thambawita et al. [2] worked on the same HyperKvasir dataset by changing the resolutions of the images to a higher resolution and observed that CNN performed significantly better, achieving a score of 0.9002 with both DenseNet-161 and Resnet-152. Additionally, both were able to classify the gastronomical organs meticulously. Following that, Sutton et al. [3] applied DenseNet121 CNN architecture on the HyperKvasir labeled images to improve the accuracy, achieving an 87.50% accuracy. The model was effective in detecting ulcerative colitis (UC) and for assessment, they employed explainable AI (Grad-Cam) for further enhanced visual interoperability.

Mohapatra et al. [4] introduced a technique fusioning Empirical Wavelet Transform (EWT) and Convolutional Neural Networks (CNNs) to detect oddities in the upper and lower gastrointestinal tract, focusing on conditions like Barrett’s, Hemorrhoids, Esophagitis, Polyps, and Ulcerative Colitis. This approach enhances feature extraction and improves CNN classification accuracy, outperforming the previous DWT [17] method. Ramamurthy et al. [5] used an advanced architecture combined with a custom-built CNN Effimix, to classify endoscopy images on the HyperKvasir dataset. Their approach enhances feature extraction, improving the differentiation of visually similar abnormalities. This model increases diagnostic accuracy while also enhancing the efficiency and reliability of gastrointestinal disease detection.

In another work, to address the issue of security and privacy concerns and ensure that sensitive data remains protected while still allowing for collaborative learning, Elbatel et al. [6] gauged how locally available self-supervised Federated Learning models can effectively enhance learning performance on the HyperKvasir dataset. To tackle the scarcity of labeled images, Guo et al. [7] utilized the unlabeled HyperKvasir dataset to enhance image classification in gastrointestinal endoscopy through self-supervised learning. They demonstrated the potential of this approach, along with data augmentation techniques C-Mixup, to improve medical image analysis and diagnosis by leveraging unlabeled data and replicating human learning strategies. Sivari et al. [8] leveraged the HyperKvasir dataset to develop novel hybrid bi-level stacking ensemble models for classifying findings in the gastrointestinal system, with a focus on facilitating early diagnosis. This approach enhances the outcomes of DL models and surpasses existing state-of-the-art models.

Nouman et al. [9] utilized a genetic algorithm to ensure optimal brightness-control contrast enhancement, improving WCE image classification accuracy by 15.26% over state-of-the-art techniques. Varam et al. [10] enhanced reliability and efficiency in endoscopy diagnosis using the Kvasir-capsule dataset. Mukhtorov et al. [11] applied explainable AI with deep learning on Kvasir images, while Khan et al. [12] extracted deep features for classification. Xu et al. [13] used a spatiotemporal STFT model on Anhui Medical University’s dataset for polyp detection. Maurício et al. [14] used deep learning models to differentiate between Crohn’s disease and ulcerative colitis by analyzing data from multiple datasets.

## III. Dataet

HyperKvasir, a large dataset for gastrointestinal endoscopy from 2020 [1], includes four main types of data: segmented images, labeled images, unlabeled images, and labeled videos. The labeled images are divided into “Upper GI Tract” and “Lower GI Tract.” Our research focused on the “Lower GI Tract,” using 7,220 labeled images gathered during real-world gastroenterological and colonoscopy procedures at Bærum Hospital which is in Norway, with some labels provided by expert endoscopists.

The primary objective of this research is to forecast and classify infected regions in the lower gastrointestinal (GI) tract. The chosen dataset helps train the model to speed up disease identification. While the upper GI tract is crucial for starting digestion, the lower GI tract plays a key role in nutrient absorption and waste processing, making its health vital. Disorders in the lower GI tract can cause serious issues like discomfort and increased cancer risk. Due to its complex anatomy and vague symptoms, this area was prioritized for focused study and treatment.

The “Lower GI Tract” dataset consists of 16 classes, divided into 4 main classes and 12 subclasses. However, CNN models struggle with small datasets, and some classes in the HyperKvasir dataset have limited data. To address this, we extracted 5 subclasses—ileum, hemorrhoids, ulcerative colitis grades 0-1, 1-2, and 2-3—using a total of 7,131 images. After excluding these 5 sub-classes, the performance and accuracy of the remaining 11 classes improved significantly compared to the original 16 classes. The distribution of classes and the number of images, both prior to and following the removal of subclasses, are presented in Table I and Table II and can be visualized in Fig 1. The dataset was initially stored in an array with corresponding labels. To improve processing, it was resized to 224×224×3, converted into a NumPy array, normalized by dividing by 255.0, and the class labels were converted from categorical to numerical values using “Label Encoder.” Categorical variables were then transformed into binary vectors with “OneHotEncoded.” The data was split into training (70%), validation (10%), and testing (20%) portions using stratified sampling to maintain class distribution. All the mentioned models, pre-trained on ImageNet, were used with the “Adam” optimizer. “Categorical crossentropy” was used as the loss metric for multiclass classification.

**TABLE I.**
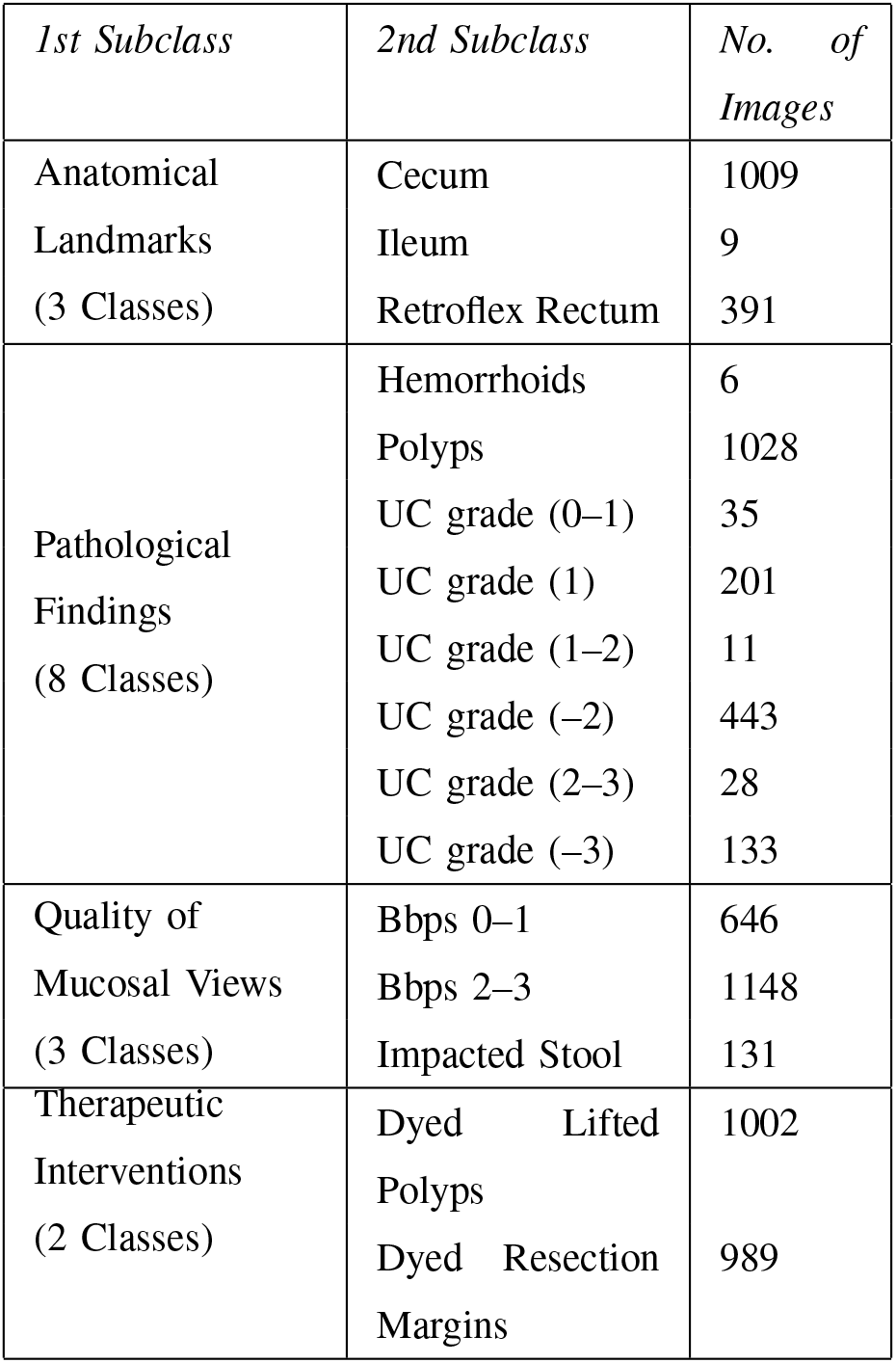
Content of the HyperKvasir Lower-GI-Tract dataset (UC = Ulcerative colitis)

**TABLE II.**
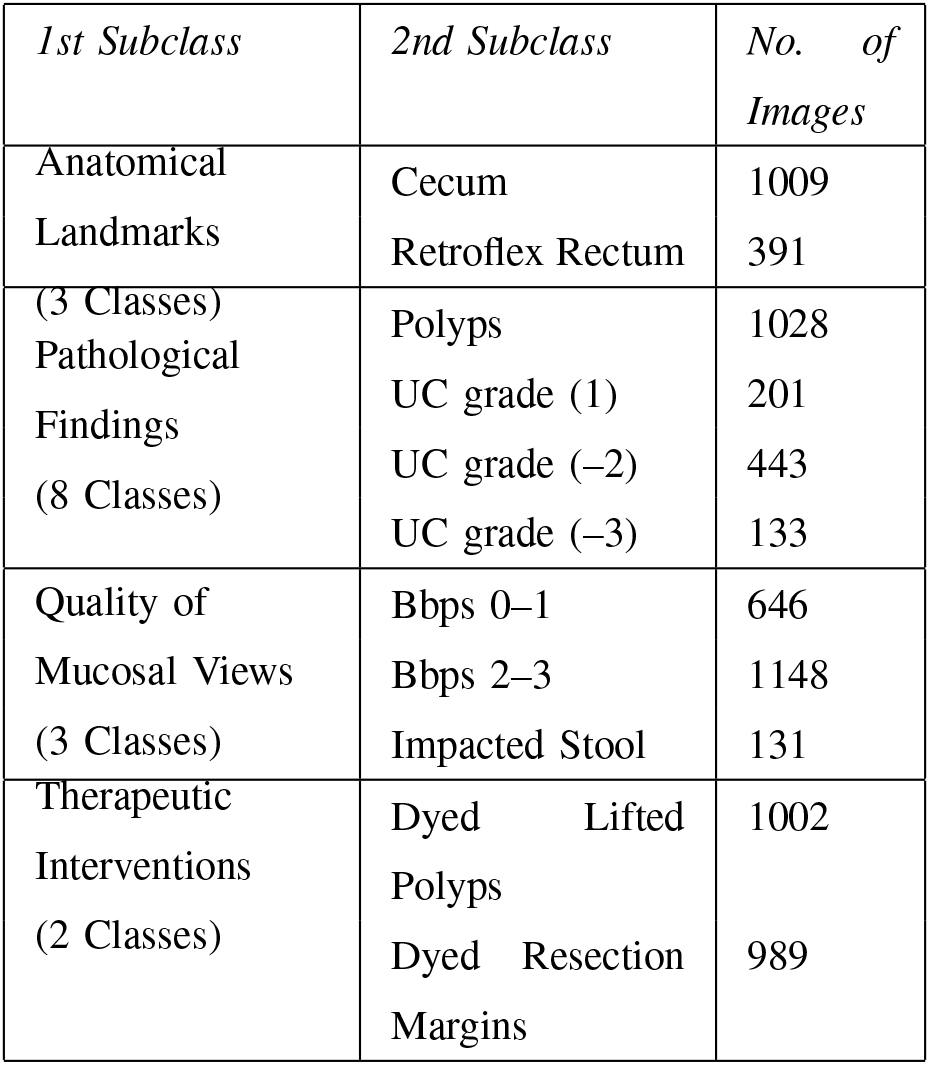
Content of the HyperKvasir Lower-GI-Tract dataset after extracting 5 subclasses (UC = Ulcerative colitis)

**Fig. 1.**
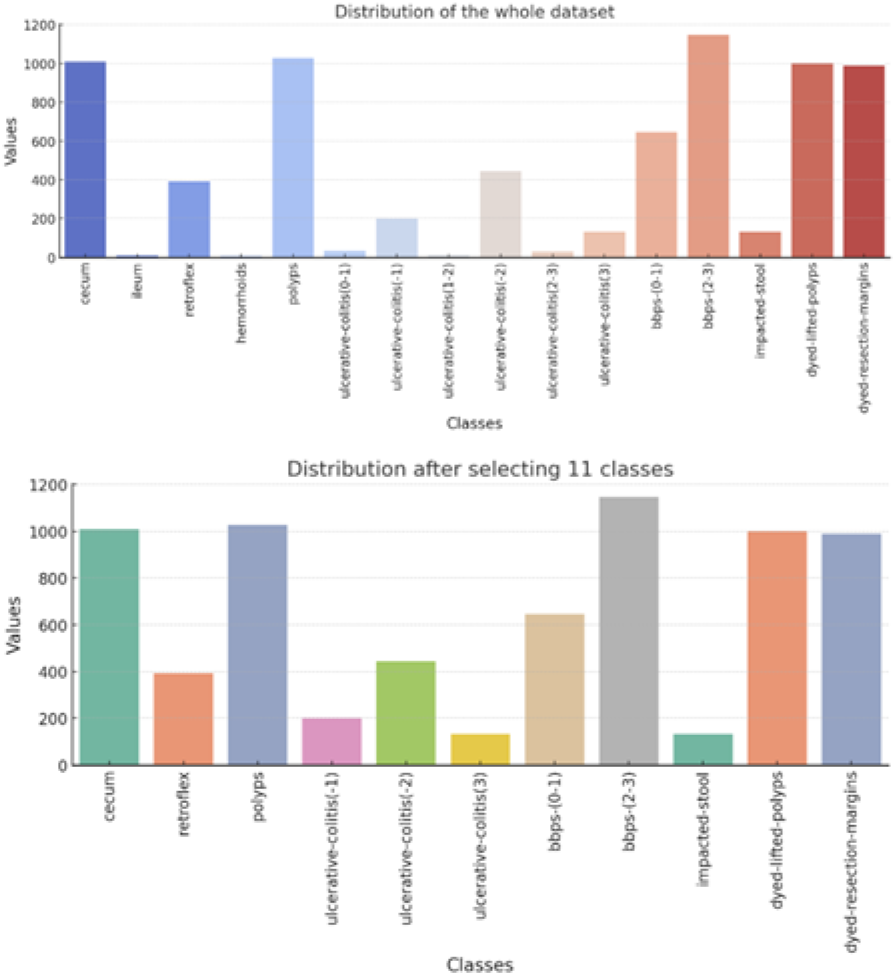
Data Distribution.

## IV. Methodology

This section outlines the pipeline for classifying gastrointestinal diseases using the HyperKvasir dataset [1]. Fig 2 visualizes the entire workflow. It includes data processing, model selection, training, evaluation, boosting algorithms and applying XAI. The dataset, consisting of 7,220 images across 16 classes, was reduced to 7,131 images across 11 classes for better performance. Collected from real procedures at Bærum Hospital, it was split into 70% training, 10% validation, and 20% testing. Five pre-trained models —VGG19 [19], ResNet50 [20], DenseNet201 [21], EfficientNetV2S [22], and InceptionNetV3 [23]—were analyzed, trained with the adam optimizer and categorical crossentropy. Training spanned 50 epochs, using early stopping and model checkpoints to optimize performance.

**Fig. 2.**
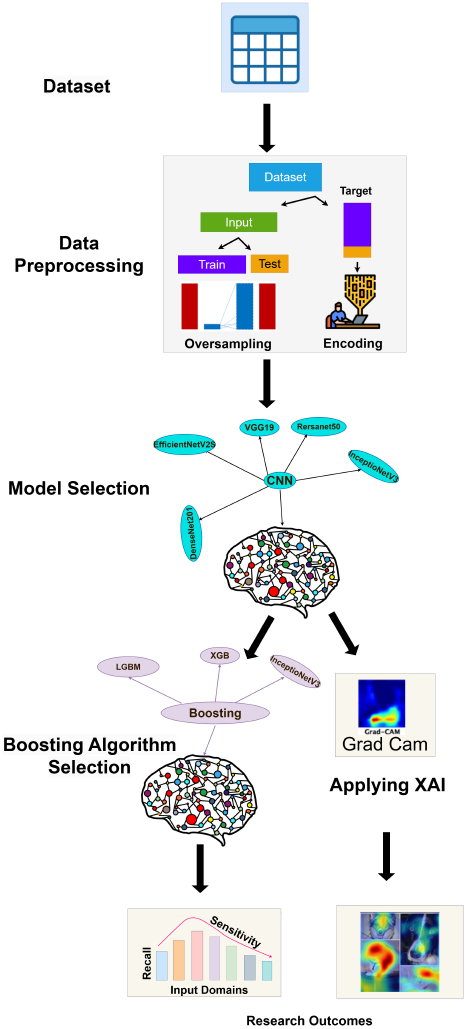
Complete Workflow.

The effectiveness of the top-performing models was evaluated using various metrics, including classification reports, confusion matrices, and training-validation-testing curves. Based results, the InceptionNetV3 model was selected for further enhancement with boosting algorithms like LGBM, XGB, and CATBOOST. Additionally, the Explainable AI (XAI) [3] method, Grad-CAM, was used to visualize the model’s decision-making by generating heatmaps that highlight the image regions most influential in predictions. This approach aims to increase accuracy and provide a clear comparison of CNN architectures in image classification tasks.

## V. Result and Performance Analysis

The performance metrics in Table III & IV are crucial for evaluating the effectiveness of different classification methods or models. We use accuracy, F1 score, precision, and recall to assess our approaches. Both train and test accuracies are shown to highlight how each model performs during the training and testing phases.

**TABLE III.**
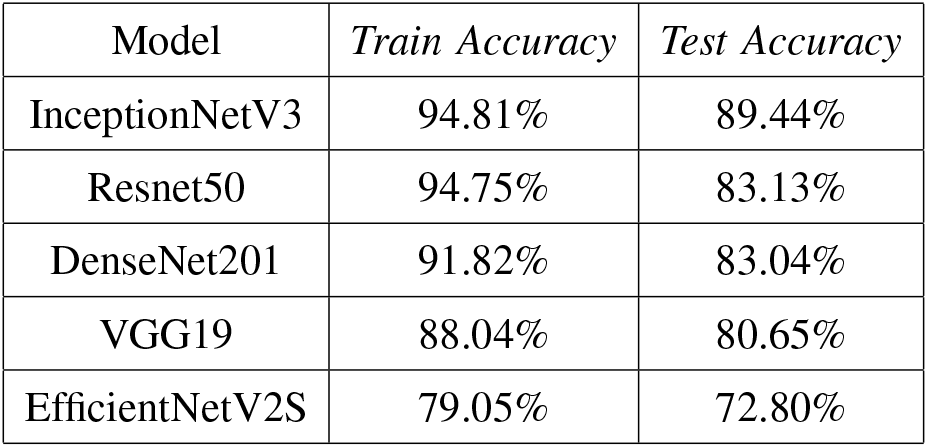
Illustration of classification report with train and test accuracy for all the models

**TABLE IV.** Illustration of classification report with Precision, Recall and F1 Score for all the models

| Model | Precision | Recall | F1-Score |
| --- | --- | --- | --- |
| InceptionNetV3 | 0.82 | 0.80 | 0.89 |
| Resnet50 | 0.76 | 0.73 | 0.83 |
| DenseNet201 | 0.76 | 0.73 | 0.83 |
| VGG19 | 0.73 | 0.73 | 0.81 |
| EfficientNetV2S | 0.76 | 0.62 | 0.73 |

First, Among the models listed in Table III & IV, InceptionNetV3 gained the peak performance across all metrics, with a training accuracy of 94.81%, test accuracy of 89.44%, precision of 0.82, recall of 0.80, and an F1-score of 0.89 in the classification task. For a fair comparison, ResNet50 and DenseNet201 demonstrated similar performance, with test accuracies around 83% and F1 scores of 0.83, showing a balanced performance between training (ResNet50 at 94.75% and DenseNet201 at 91.82%) and testing, though slightly trailing InceptionNetV3. In contrast, VGG19 exhibited lower training accuracy at 88.04%, reflecting weaker performance during training. Its test accuracy of 80.65% indicates good generalization, with precision and recall of 0.73 and an F1 score of 0.81, offering balanced but modest classification performance.

However, EfficientNetV2S had the lowest training accuracy at 79.05% and test accuracy at 72.80%, demonstrating poor generalization. Despite a relatively good precision of 0.76, its recall was lower at 0.62, resulting in the lowest F1 score of 0.73, highlighting a significant imbalance between precision and recall in contrast to the other models.

The table V provides a comparison of classification metrics (Precision, Recall, F1 Score) for various deep learning models (InceptionNetV3, ResNet50, DenseNet201, VGG19, EfficientNetV2S) when combined with three boosting algorithms (LGBM, XGB, CATBOOST).

**TABLE V.** Illustration of classification report of boosting algorithm’sS Precision, Recall and F1 Score accuracy for all the models

| Model | Precision | Recall | F1-Score |
| --- | --- | --- | --- |
| (InceptionNetV3) |  |  |  |
| LGBM | 0.80 | 0.80 | 0.89 |
| XGB | 0.81 | 0.81 | 0.90 |
| CATBOOST | 0.81 | 0.81 | 0.89 |
| (Resnet50) |  |  |  |
| LGBM | 0.79 | 0.75 | 0.86 |
| XGB | 0.79 | 0.76 | 0.86 |
| CATBOOST | 0.77 | 0.75 | 0.85 |
| (DenseNet201) |  |  |  |
| LGBM | 0.77 | 0.74 | 0.84 |
| XGB | 0.79 | 0.76 | 0.85 |
| CATBOOST | 0.74 | 0.72 | 0.83 |
| (VGG19) |  |  |  |
| LGBM | 0.77 | 0.71 | 0.83 |
| XGB | 0.75 | 0.72 | 0.83 |
| CATBOOST | 0.72 | 0.71 | 0.82 |
| (EfficientNetV2S) |  |  |  |
| LGBM | 0.80 | 0.79 | 0.88 |
| XGB | 0.81 | 0.80 | 0.89 |
| CATBOOST | 0.82 | 0.81 | 0.88 |

InceptionNetV3 combined with XGB in Table V demonstrated the best performance across all metrics, achieving a Precision of 0.81, Recall of 0.81, and an F1 Score of 0.90. This combination proved to be the most optimal, outperforming other boosting methods in terms of overall classification performance. The EfficientNetV2S model also showed strong results with XGB, achieving a Precision of 0.81, Recall of 0.80, and an F1 Score of 0.89. In particular, it performed well with CATBOOST, where it recorded an F1 Score of 0.88 along with high Precision and Recall. This model provides a competitive alternative. ResNet50 and DenseNet201 models showed close F1 Score accuracy compared to InceptionNetV3 and EfficientNetV2S, with values 0.86 and 0.85 across the XGB boosting algorithms, indicating moderate performance but less than InceptionNetV3 XGB. VGG19 performed the lowest with XGB, achieving an F1 Score of 0.83. and it remains the least effective among the models tested.

Fig. 3 illustrates the confusion matrix for the InceptionNetV3 model, which performs well in classifying 11 medical categories, particularly bbps-(0-1), bbps-(2-3), cecum, dyed-lifted polyps, dyed-resection margins, and polyps. However, some misclassifications occur between similar categories like dyed-lifted polyps and dyed-resection margins. There are also difficulties in distinguishing between ulcerative colitis grades (−1, -2, -3), especially between grades (−2) and (−3). In contrast, Fig. 4 presents the confusion matrix for the XGB model applied to InceptionNetV3, which similarly performs well but encounters the same challenges with misclassi-fying ulcerative colitis grades.

**Fig. 3.**
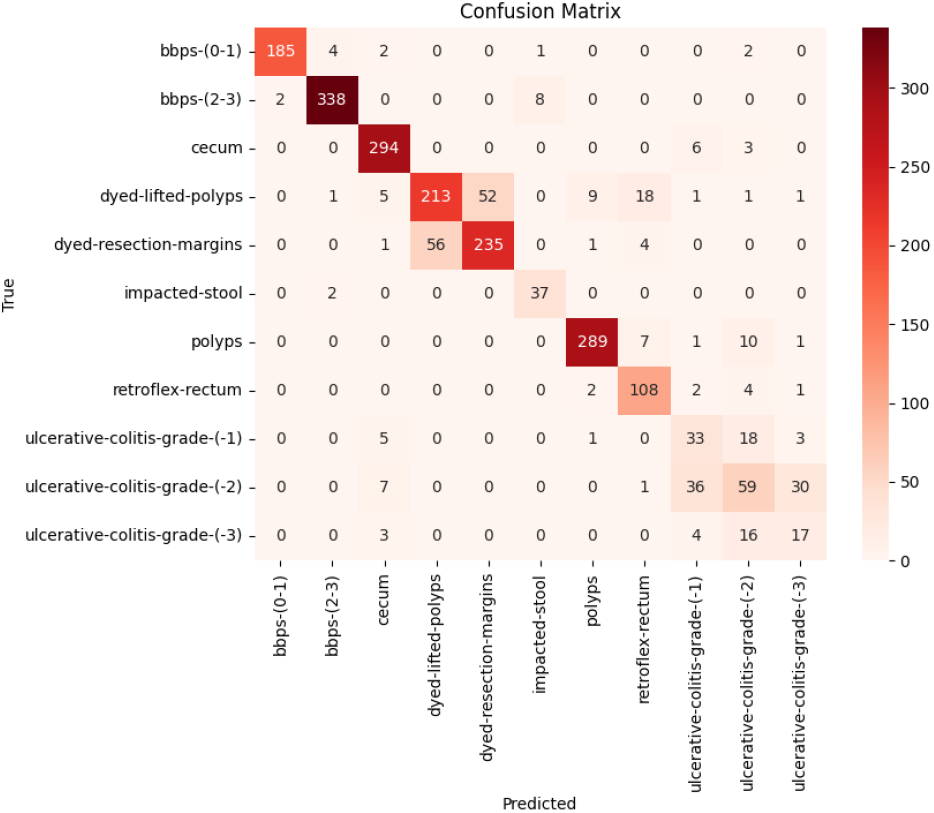
Confusion Matrix of Pre-trained InceptionNetV3.

**Fig. 4.**
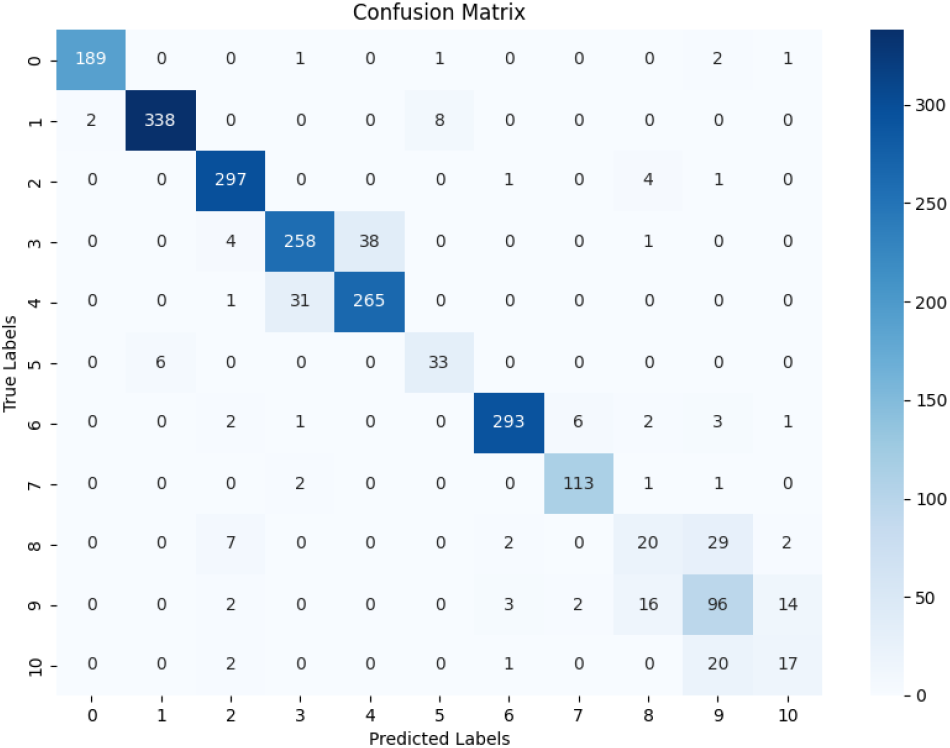
Confusion Matrix of XGB on Pre-trained InceptionNetV3.

## VI. Explainable AI (XAI)

Explainable AI (XAI) has developed into a multidisciplinary area, driven by the increasing application of AI in sectors such as finance and healthcare. As Machine Learning (ML) and Deep Learning (DL) continue to be widely used for tasks like classification and prediction, it becomes crucial to understand the typically opaque inner workings of these models. XAI addresses this issue by developing methods to clarify AI systems and their outputs, fostering transparency and accountability, particularly in critical domains. Specifically, XAI [18] enhances ML classification by illuminating the decision-making processes of models, identifying significant factors that impact results, and building user trust. This heightened transparency improves model interpretability, aids in debugging, uncovers biases, and enhances model performance. Grad-CAM is one such visualization technique that highlights key image regions that influence a model’s class predictions, especially in CNNs. It is valuable in image classification, object detection, and medical imaging, facilitating the interpretation of model decisions. The heatmaps, which represent the gradient-weighted class activations for each of the three classification tasks, were produced using the InceptionNetV3 architecture and are presented in Fig. 5

**Fig. 5.**
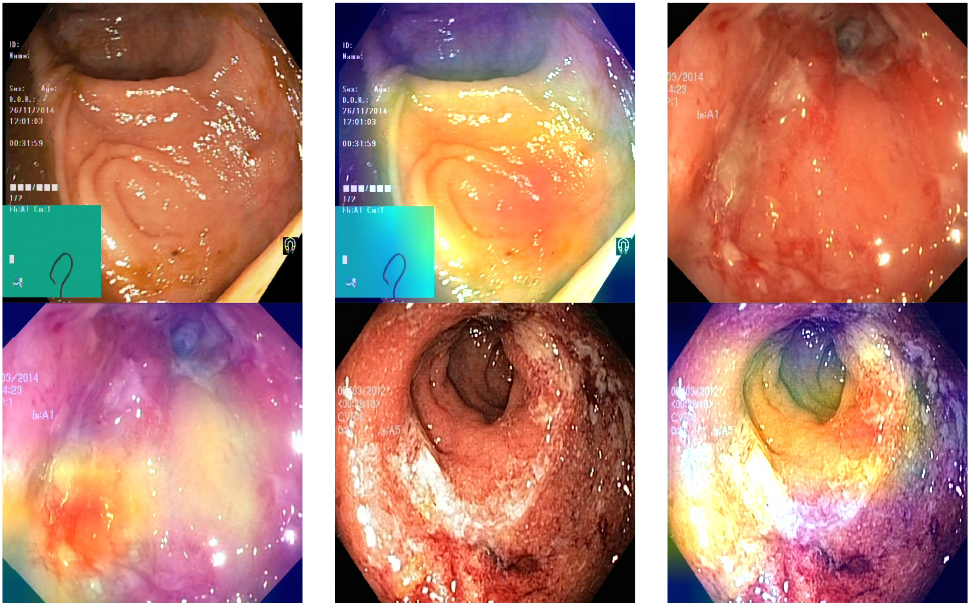
Class activation heatmaps alongside the original endoscopic images.

## VII. Conclusion

This research shows that deep learning models and boosting algorithms can effectively classify gastrointestinal (GI) images, helping to diagnose GI diseases more accurately. Using the HyperKvasir dataset, models such as InceptionNetV3, ResNet50, DenseNet201, VGG19, and EfficientNetV2S, with InceptionNetV3 achieving an F1-Score of 0.89. Moreover, combining InceptionNetV3 with the XGB boosting method yielded the best result, with a peak F1 score of 0.90, which outperforms the other state-of-the-art model work.

To make these models more trustworthy for doctors, the Grad-CAM technique was used. This method highlights the important areas in images that the models focus on when making decisions, making the results more understandable. Even though GI images vary a lot, this study proves that boosting algorithms and explainable AI (XAI) can improve the diagnosis process. These methods can help to reduce errors and provide quicker, more accurate results in medical settings. In the future, improving these models and making them easier to use in real medical environments will be important steps forward. This research contributes to making automated systems more reliable for diagnosing GI diseases, ultimately benefiting patient care.

Bridging the gaps, Future work should focus on expanding dataset diversity, particularly in underrepresented classes, enhancing the interpretability of Grad-CAM, and improving feature extraction to address misclassifications. Developing lightweight models, validating them in clinical settings, and extending the scope to include the upper GI tract are essential. Incorporating multimodal data, utilizing few-shot learning, and exploring federated learning can further improve accuracy, security, and practical applicability.

## Data Availability

All data produced in the present study are available upon reasonable request to the authors

